# Pharmacokinetics and Pharmacodynamics of Oral and Vaporized Δ9-Tetrahydrocannabinol in Older Adults

**DOI:** 10.64898/2026.08.18.26360706

**Authors:** Gabriel P. A. Costa, Simon Asnes, Julia Meyerovich, Tore Eid, Haleh Nadim, Stephanie Dwy, Ralitza Gueorguieva, Matthew M. Riggs, Mehmet Sofuoglu, Scott Matthews, Julio C. Nunes, Joao P. De Aquino

## Abstract

Adults aged ≥65 years are increasingly using cannabis products. However, controlled pharmacokinetic and pharmacodynamic data on Δ9-tetrahydrocannabinol (THC) in this population are sparse, and remain limited to oral/oromucosal formulations. To characterize the acute pharmacokinetic and pharmacodynamic effects of oral and vaporized THC in healthy adults aged ≥65, we conducted a two-arm, randomized, double-blind, placebo-controlled trial in which 20 participants (mean age 70.0, SD: 5.1 years) received oral (placebo, 5 mg, or 10 mg) or vaporized THC (placebo, 2 mg, or 4 mg) across three eight-hour sessions separated by ≥72 hours. Outcomes included plasma pharmacokinetics, subjective drug effects, reinforcement value, cognitive performance, heart rate (HR), blood pressure (BP), and adverse events (AEs). Oral THC was associated with delayed, lower THC exposure (Tmax 60-90 min; Cmax 2.6-6.2 ng/mL), with 11-OH-THC concentrations approximately matching parent-THC; slow-rising subjective effects; no change in reinforcement value; no significant change in HR or BP; and no AEs. Vaporized THC was associated with rapid, THC-dominant exposure (Tmax 3 min; Cmax 24.6-53.8 ng/mL) and minimal 11-OH-THC concentrations; rapid-onset subjective effects; increased reinforcement value at 4 mg; and significant HR elevation peaking within 5 min, without significant BP change. Cognitive performance did not differ from placebo at any oral or vaporized THC dose. At vaporized THC 4 mg, two participants experienced five AEs. Oral and vaporized THC produce route-specific pharmacokinetic and pharmacodynamic profiles in adults aged ≥65, including an increase in reinforcement value only after vaporization, and should therefore not be treated as interchangeable in risk assessment for older adults.

## 1. Introduction

Cannabis use is rising faster among older adults than in any other U.S. age group. Over the past two decades, adults aged 65 and older have increased their past-year cannabis use 20-fold[1–5], while their perceived risk of cannabis use declined by 20% [6]. Over the same period, clinical interest in cannabinoid pharmacotherapy has expanded for disorders common among older adults, including chronic pain, sleep disorders, and behavioral symptoms of dementia[7–10].

Cannabis’s acute effects are primarily produced by Δ9-tetrahydrocannabinol (THC), its principal psychoactive constituent[11], which acts as a partial agonist at cannabinoid 1 (CB1) receptors[12] and produces euphoria, reinforcing effects, impaired attention and memory, and increased heart rate[13–16]. Each of these effects carries clinical weight in older adults: the subjective and reinforcing effects are relevant to the risk of cannabis use disorder, which affects roughly one in three older adults who use cannabis[2], while the cognitive and cardiovascular effects may intersect with age-related vulnerabilities. Notably, findings from younger cohorts cannot be directly extrapolated to older adults, as aging alters both the pharmacodynamics and pharmacokinetics of THC [17].

At the pharmacodynamic level, CB1 receptor density declines with age, changing the substrate through which THC produces its cognitive and cardiovascular effects[18,19]. Cognitively, adults aged 55-70 showed less acute impairment in learning and processing speed than adults aged 21-25 given comparable exposure[20]. Cardiovascular reserve and autonomic regulation also diminish with age, increasing the potential consequences of THC-related tachycardia and orthostatic hypotension[14,21–23], including an elevated risk of falls[24,25].

At the pharmacokinetic level, THC is metabolized in the liver, primarily by CYP2C9, to the active metabolite 11-hydroxy-THC (11-OH-THC) and then to the inactive 11-nor-9-carboxy-THC (THC-COOH)[26–30]; greater adiposity may expand THC’s volume of distribution[31,32], and reduced hepatic blood flow may lower clearance.

Route of administration is another determinant of THC exposure. Oral routes, including edibles, represent a growing share of cannabis use[33–35]. Inhalation, historically dominated by combustion but increasingly delivered through vaporization, accounts for most non-medical use[33,35]. While oral THC administration leads to conversion to the active metabolite 11-OH-THC, most of the effects of vaporized THC are attributed to THC itself[26,36,37]. Whether these route-specific profiles are maintained in older adults or reshaped by the age-related changes in distribution and clearance has not been investigated.

Despite the convergence of epidemiologic, clinical, pharmacodynamic, and pharmacokinetic rationales, controlled human laboratory evidence in older adults remains sparse. Two trials examined Namisol, an oromucosal tablet of purified THC[38,39], and two trials examined dronabinol or nabilone for agitation in participants with dementia[9,40]. Thus far, no study has characterized the effects of vaporized THC in older adults, or compared abuse liability, cognitive performance, and cardiovascular response across routes in this population.

Therefore, to characterize the acute pharmacokinetic and pharmacodynamic effects of THC in healthy adults aged ≥65, we conducted a randomized, double-blind, placebo-controlled clinical trial. Participants received THC through the two routes most clinically relevant in this population: oral and vaporized. Across both routes, we measured plasma pharmacokinetics, abuse liability (subjective drug effects and reinforcement value), cognitive performance, cardiovascular effects, and tolerability. We administered THC rather than cannabis because cannabinoid products are standardized and dosed by their THC content[41], and because isolating THC affords precise characterization of its phase I and phase II metabolism. These results are intended as a benchmark extensible to full-spectrum products, other cannabinoids, and clinical populations.

## 2. Methods

This study was registered on ClinicalTrials.gov (NCT05906511) and conducted at the Biological Studies Unit of the VA Connecticut Healthcare System (West Haven, CT). The protocol was approved by Yale University and the VA Connecticut Healthcare System Institutional Review Boards. All participants provided written informed consent before study procedures. Reporting follows the Consolidated Standards of Reporting Trials (CONSORT) 2025 guidelines[42].

### 2.1 Study Design

This was a two-arm, randomized, double-blind, placebo-controlled trial. Participants were assigned to either an oral or a vaporized arm and, within their assigned arm, completed three eight-hour test sessions in randomized counterbalanced order: placebo, 5 mg, or 10 mg of THC in the oral arm; placebo, 2 mg, or 4 mg of THC in the vaporized arm. Sessions were separated by a minimum 72-hour washout period to minimize carry-over effects.

### 2.2 Participants

Eligible participants were healthy adults aged ≥65 with documented prior cannabis exposure (≥1 exposure in the last 10 years; 1-10 times in the last 20 years; or ≥20 times in their lifetime) to enhance safety and tolerability. History of cannabis use disorder was excluded to minimize tolerance, residual cannabinoid exposure, and withdrawal. Participants were required to provide a negative urine sample for THC-COOH (≤50 ng/mL) prior to drug administration at each test session.

Additional exclusion criteria covered past-year DSM-5 substance use disorders other than tobacco use disorder, clinically significant hepatic, renal, or immunosuppressing conditions, personal or family history of psychotic spectrum disorders, allergy to sesame oil or THC, and concomitant medications that interact with THC metabolism. Full criteria are listed in **Section S1**.

### 2.3 Study Medication

Oral THC was administered as dronabinol (synthetic THC), an FDA-approved Schedule III oral formulation with bioavailability of approximately 20%[29,30,43]. Participants received visually identical capsules containing 0, 5, or 10 mg of THC, comparable to the lower range of doses prescribed in clinical practice[44] and to those used in the only published controlled trial in older adults[38,39].

Vaporized THC was delivered using the Volcano Hybrid® vaporizer (Storz & Bickel), validated for reproducible, smoke-free cannabinoid delivery[45], using pharmaceutical-grade THC (99% purity) or placebo (ethanol), inhaled following a standardized puffing procedure[46,47]. Vaporized doses of 2 and 4 mg were selected to approximate the systemic exposure of the 5 and 10 mg oral doses, reflecting the greater bioavailability of inhaled THC[48] and keeping exposure within the low-to-moderate range of prior human laboratory studies[49–51]. Detailed preparation procedures are provided in **Section S2**.

To assess blinding, participants and research staff independently guessed the assigned condition after each session. Blinding was summarized for each treatment condition as percent correct and Bang’s Blinding Index[52].

### 2.4 Pharmacokinetic Assessments

Plasma was sampled via an indwelling intravenous catheter at 12 protocol-specified timepoints per session: a pre-dose baseline, 10 post-dose timepoints, and a 24-hour follow-up sample. The post-dose schedule was arm-specific: dense mid-session sampling in the oral arm (Tmax 60-90 min) and dense early sampling in the vaporized arm (Tmax ∼3 min). The full schedule is provided in **Table S1**. Plasma concentrations of THC, 11-OH-THC, and THC-COOH were quantified by liquid chromatography-tandem mass spectrometry (LC-MS/MS), with lower limits of quantification of 0.2 ng/mL for THC, 1.0 ng/mL for 11-OH-THC, and 0.5 ng/mL for THC-COOH.

### 2.5 Subjective Effects

We assessed subjective drug effects using the Drug Effects Questionnaire (DEQ), a self-report, 10-item visual analog scale (0-100 mm) with established psychometric properties across drug classes[53] and demonstrated sensitivity to acute effects of THC in humans[54,55], in line with FDA guidance for assessing abuse liability[56].

Consistent with prior studies, we averaged the items into three composite subscales: *Stimulatory* (feel drug effect, feel high, feel stimulated), *Pleasurable* (like drug effects, feel good effects, want more drug), and *Aversive* (feel anxious, feel down or sad, feel bad effects)[57–59]. The DEQ was administered at a pre-dose baseline and at protocol-specified post-dose timepoints (14 in the oral arm, 13 in the vaporized arm).

### 2.6 Reinforcement Value

As a complement to the DEQ, we used the Multiple-Choice Procedure (MCP)[60], a validated analog drug self-administration assessment that is sensitive to the effects of THC, as an index of abuse liability[61–63].

At the end of each session, participants made a series of hypothetical choices between receiving the just-completed session’s dose again or escalating dollar amounts. Consistent with previous studies[61–63], the crossover point — the lowest dollar value at which the participant first preferred money over the dose — served as the session-level reinforcement value.

### 2.7 Cognitive Performance

We assessed sustained attention with the Continuous Performance Test (CPT)[64], using throughput (combined accuracy and reaction time) as the primary outcome. We assessed verbal learning and memory with the Hopkins Verbal Learning Test-Revised (HVLT-R)[65], using Total Recall (sum across three learning trials of a 12-word list) and Delayed Recall (after a 20-minute delay). Both tasks are sensitive to acute THC exposure[13,15], and the HVLT-R was used in prior cannabinoid trials in adults aged ≥65[38,39]. Testing occurred once per session, timed to the expected period of prominent subjective drug effects: 150 and 30 minutes after oral and vaporized dosing, respectively.

### 2.8 Heart Rate and Blood Pressure

Heart rate (HR), systolic blood pressure (SBP), and diastolic blood pressure (DBP) were measured with an automated oscillometric monitor. Measurements were taken at 17 protocol-specified timepoints per session: one pre-dose baseline, 15 post-dose timepoints, and one 24-hour follow-up.

### 2.9 Adverse Events

We assessed safety and tolerability by monitoring treatment-emergent symptoms with the Systematic Assessment for Treatment-Emergent Events (SAFTEE), a standardized instrument widely used in human laboratory trials of psychoactive drugs[66]. The SAFTEE was administered by trained research staff at a pre-dose baseline and at discharge (480 minutes after drug administration). An adverse event was counted whenever a SAFTEE symptom was rated higher in severity at discharge than at that session’s pre-dose baseline.

### 2.10 Statistical Analysis

Pharmacokinetic parameters, including area under the curve from 0 to 8 hours (AUC0-8h), maximum plasma concentration (Cmax), and time to maximum concentration (Tmax), were derived by non-compartmental analysis using the *PKNCA* package[67]. Between-subject variability is reported as the coefficient of variation (CV=standard deviation[SD]/mean × 100%). Exploratory associations of Cmax and AUC0-8h with body weight are reported in **Section S17**.

Pharmacodynamic outcomes were analyzed with linear mixed models using the mmrm package[68], including dose and session as fixed effects, and dose as a within-participant repeated factor, with heterogeneous compound-symmetry structure, allowing response variability to differ by dose. Denominator degrees of freedom used the Kenward-Roger approximation. For DEQ, HR, and BP, we analyzed the peak change from baseline over the session: the maximum increase for DEQ and HR, and the maximum decrease for SBP and DBP. This follows FDA abuse-liability guidance for DEQ[56] and prior controlled cannabinoid studies for HR and BP[50]. Cognitive performance assessments and the MCP were modeled identically. We report pairwise dose contrasts as mean differences (MD) with 95% confidence intervals in the outcome’s original units, accompanied by Hedges’ g, computed from the model-estimated dose-specific SDs[69].

Finally, we examined within-participant exposure–response relationships for subjective effects, HR, SBP, and DBP by relating each participant’s peak post-dose response to peak active combined analytes concentration (Cmax; THC plus 11-OH-THC) across sessions with linear mixed-effects models (**Section S16**). All analyses were performed in *R* version 4.5.2[70] with two-sided α=0.05.

## 3. Results

### 3.1 Participants

Twenty older adults were enrolled, 10 per arm (**Table 1**, **Figure S1**). Overall, mean age was 70.0 (SD: 5.1) years, 30% (n=6) were female, and mean BMI was 28.6 (SD: 4.2) kg/m². The two arms were balanced on demographic and cannabis use characteristics (all p≥.20). In the oral arm, 20% (n=2) were female, mean age was 70.5 (SD: 4.7) years, and mean BMI was 27.6 (SD: 4.2) kg/m². In the vaporized arm, 40% (n=4) were female, mean age was 69.5 (SD: 5.7) years, and mean BMI was 29.6 (SD: 4.2) kg/m².

**Table 1.** Participant Characteristics.

| Participant Characteristics |  | Treatment arm |  | p-value<sup>2</sup> |
| --- | --- | --- | --- | --- |
| Characteristic | Overall | Oral | Vaporized |  |
| **Age, years** | 70.0 (5.1) | 70.5 (4.7) | 69.5 (5.7) | 0.67 |
| **Sex (male)** | 14 (70%) | 8 (80%) | 6 (60%) | 0.63 |
| **Race** |  |  |  | **>0.99** |
| Black | 0 (0%) | 0 (0%) | 0 (0%) |  |
| Asian | 0 (0%) | 0 (0%) | 0 (0%) |  |
| Native American | 0 (0%) | 0 (0%) | 0 (0%) |  |
| White | 19 (95%) | 10 (100%) | 9 (90%) |  |
| Other | 1 (5.0%) | 0 (0%) | 1 (10%) |  |
| **Ethnicity** |  |  |  | **>0.99** |
| Hispanic | 1 (5.0%) | 0 (0%) | 1 (10%) |  |
| Not Hispanic | 19 (95%) | 10 (100%) | 9 (90%) |  |
| **Height, cm** | 171.6 (12.1) | 170.9 (16.3) | 172.2 (6.5) | 0.82 |
| **Weight, kg** | 84.5 (16.7) | 80.7 (16.5) | 88.2 (16.9) | 0.33 |
| **Body mass index, kg/m<sup>2</sup>** | 28.6 (4.2) | 27.6 (4.2) | 29.6 (4.2) | 0.31 |
| **Body fat, %<sup>3</sup>** | 30.5 (4.4) | 28.8 (2.7) | 31.8 (5.1) | 0.22 |
| **Age at first cannabis use, years** | 17.0 (2.8) | 17.2 (2.9) | 16.7 (2.9) | 0.70 |
| **Total years of cannabis use** | 33.6 (19.2) | 33.6 (19.7) | 33.7 (19.9) | 0.99 |
| **Past 30-day use (0-8 ordinal)** | 3.4 (2.9) | 3.4 (2.7) | 3.3 (3.2) | 0.94 |
| **Lifetime days of cannabis use** |  |  |  | **>0.99** |
| 0 | 0 (0%) | 0 (0%) | 0 (0%) |  |
| 1-10 | 3 (15%) | 1 (10%) | 2 (20%) |  |
| 11-50 | 0 (0%) | 0 (0%) | 0 (0%) |  |
| 51-100 | 1 (5.0%) | 1 (10%) | 0 (0%) |  |
| 101-200 | 1 (5.0%) | 0 (0%) | 1 (10%) |  |
| 201-300 | 1 (5.0%) | 1 (10%) | 0 (0%) |  |
| >300 | 14 (70%) | 7 (70%) | 7 (70%) |  |
| **Typical method of consumption** |  |  |  | **0.20** |
| Joint | 9 (47%) | 7 (70%) | 2 (22%) |  |
| Bowl | 2 (11%) | 1 (10%) | 1 (11%) |  |
| Bong | 2 (11%) | 1 (10%) | 1 (11%) |  |
| One-hitter | 2 (11%) | 0 (0%) | 2 (22%) |  |
| Ingestion (e.g. food) | 4 (21%) | 1 (10%) | 3 (33%) |  |

<sup>1</sup> Mean (SD:) for continuous variables; n (%) for categorical variables.
<sup>2</sup> Welch's two-sample t-test for continuous variables; Fisher's exact test for categorical variables. No multiplicity correction (balance check, exploratory).
<sup>3</sup> Body fat percentage was available for 12 of 20 participants (oral, 5 of 10; vaporized, 7 of 10) and is summarized on available data.

### 3.2 Pharmacokinetics

Oral and vaporized administration produced distinct pharmacokinetic profiles (**Table 2**, **Figure 1**). After oral THC, exposure was delayed and lower in magnitude (Tmax 90 and 60 minutes; mean Cmax 2.64 and 6.16 ng/mL at 5 and 10 mg), with substantial 11-OH-THC formation matching THC at both doses (11-OH-THC:THC Cmax ratio ∼1.0). THC-COOH rose more gradually, peaking at 120 minutes.

**Figure 1.**
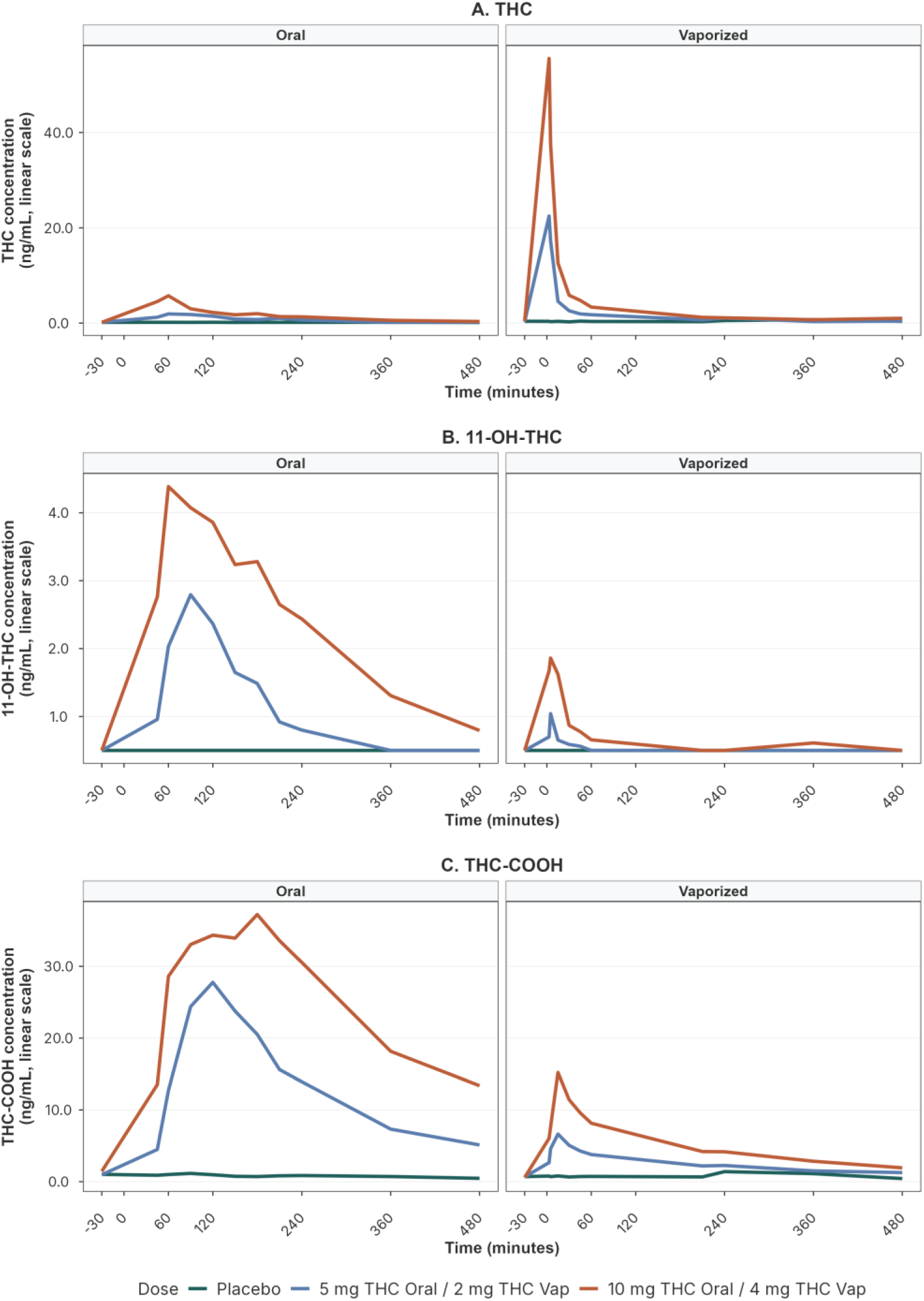
Plasma pharmacokinetics of THC and metabolites after oral and vaporized THC in healthy adults aged ≥65 years. Mean plasma concentrations of (A) Δ9-tetrahydrocannabinol (THC), (B) 11-hydroxy-Δ9-tetrahydrocannabinol (11-OH-THC), and (C) 11-nor-9-carboxy-Δ9-tetrahydrocannabinol (THC-COOH) across the 480-min observation window. Results are shown separately by route of administration: oral THC in the left panels and vaporized THC in the right panels. Conditions are shown as placebo (green), 5 mg oral / 2 mg vaporized THC (blue), and 10 mg oral / 4 mg vaporized THC (orange). Concentrations are plotted on a linear scale.

**Table 2.** Pharmacokinetic parameters of THC and metabolites after oral and vaporized administration in healthy adults aged ≥65 years.

| Pharmacokinetic Parameters by Route and Dose |  |  |  |  |  |  |
| --- | --- | --- | --- | --- | --- | --- |
| Route | Dose | Analyte | C<sub>max</sub>, mean (SD), ng/mL | T<sub>max</sub>, min | AUC<sub>0-8h</sub>, ng·h/mL | CV% |
| Oral | 5 mg | THC | 2.64 (1.93) | 90 | 5.07 | 73.3 |
|  |  | 11-OH-THC | 2.77 (2.01) | 90 | 7.66 | 72.7 |
|  |  | THC-COOH | 27.33 (11.48) | 120 | 95.26 | 42.0 |
|  | 10 mg | THC | 6.16 (3.77) | 60 | 12.92 | 61.2 |
|  |  | 11-OH-THC | 6.23 (2.44) | 105 | 18.78 | 39.1 |
|  |  | THC-COOH | 56.98 (21.34) | 120 | 194.97 | 37.4 |
| Vaporized | 2 mg | THC | 24.55 (13.74) | 3 | 9.68 | 56.0 |
|  |  | 11-OH-THC | 0.72 (1.12) | 5 | 1.11 | 154.9 |
|  |  | THC-COOH | 6.73 (2.64) | 15 | 19.53 | 39.3 |
|  | 4 mg | THC | 53.76 (25.18) | 3 | 17.90 | 46.8 |
|  |  | 11-OH-THC | 1.93 (0.94) | 5 | 3.60 | 48.9 |
|  |  | THC-COOH | 12.78 (5.35) | 15 | 31.68 | 41.8 |

After vaporized THC, exposure was more rapid and higher: Tmax was 3 minutes at both doses, and peak concentrations were approximately 9-fold higher than at the corresponding oral doses and occurred 20- to 30-fold faster, with total exposure (AUC0-8h) 1.4- to 1.9-fold higher. In contrast to oral THC, 11-OH-THC exposure was minimal (11-OH-THC:THC Cmax ratio ∼0.03-0.04), and THC-COOH was roughly 4-fold lower at peak and 5- to 6-fold lower in total exposure. Interindividual variability was substantial across analytes and routes, particularly for 11-OH-THC at 2 mg vaporized THC (**Figure S2**). Greater body weight was inversely associated with THC Cmax (p=.021), 11-OH-THC Cmax (p=.018), and 11-OH-THC AUC0-8h (p=.049) at the 10 mg oral dose (**Section S17**, **Table S9**).

### 3.3 Subjective Effects

Subjective effects mirrored each route’s pharmacokinetic profile over time (**Figure 2A-C**). In the oral arm, peak Pleasurable and Stimulatory ratings differed by dose (p=.038 and .004, respectively). Both doses of oral THC exceeded placebo, such that ratings peaked between 90 and 150 min. At peak, Pleasurable ratings were higher than placebo by 19.1 points at 5 mg (p=.021, g=0.84) and 22.8 points at 10 mg (p=.034, g=0.77), while Stimulatory ratings were higher by 25.3 (p=.007, g=1.19) and 33.3 points (p=.002, g=1.41), respectively (**Table 3**). Aversive ratings exceeded placebo at 5 mg by 6.6 points (p=.046, g=0.68).

**Figure 2.**
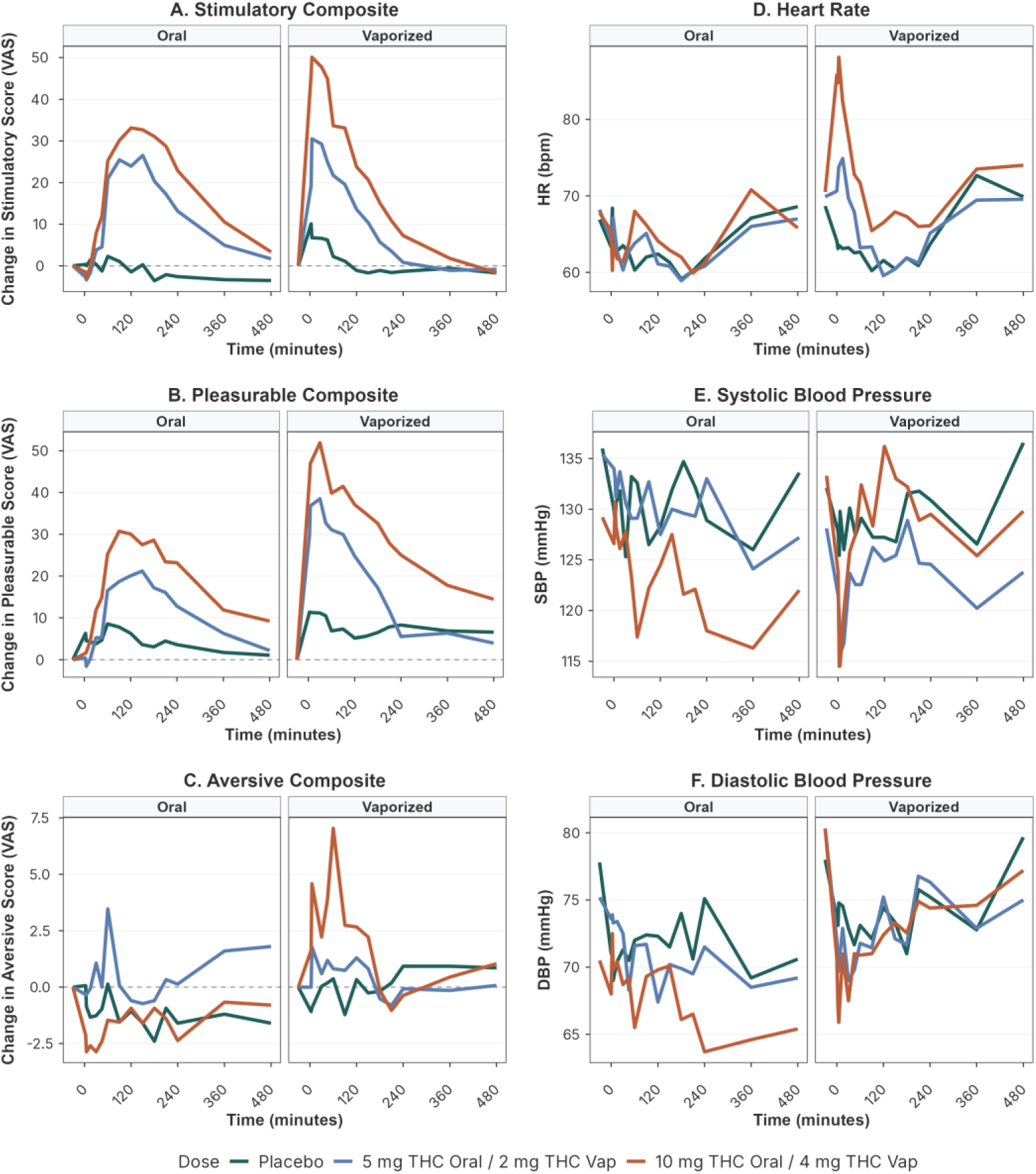
Time course of subjective drug effects, heart rate, and blood pressure after oral and vaporized THC in healthy adults aged ≥65 years. Mean change from baseline on the three Drug Effects Questionnaire (DEQ) composites: (A) Stimulatory (mean of feel drug effect, feel high, feel stimulated), (B) Pleasurable (mean of like drug effects, feel good effects, want more drug), and (C) Aversive (mean of feel anxious, feel down or sad, feel bad effects); and mean (D) heart rate (HR, bpm), (E) systolic blood pressure (SBP, mmHg), and (F) diastolic blood pressure (DBP, mmHg), across the 480-min observation window. Results are shown separately by route of administration: oral THC in the left panels and vaporized THC in the right panels. Conditions are shown as placebo (green), 5 mg oral / 2 mg vaporized THC (blue), and 10 mg oral / 4 mg vaporized THC (orange). DEQ items were rated on 0-100 mm visual analog scales; the dashed horizontal line marks no change from baseline.Panels A-C show change from baseline; panels D-F show absolute values, obtained by automated oscillometric monitor.

**Table 3.** Significant pharmacodynamic effects by route and dose (peak effect vs placebo).

| Pharmacodynamic Effects by Route and Dose |  |  |  |  |  |  |  |
| --- | --- | --- | --- | --- | --- | --- | --- |
| Domain | Outcome | Route | Dose | Peak time | MD [95% CI] | Hedges' g | p |
| Subjective | Pleasurable (VAS) | Oral | 5 mg | 150 min | +19.1 [+3.4, +34.8] | +0.84 | .021 |
|  |  |  | 10 mg | 90 min | +22.8 [+2.0, +43.6] | +0.77 | .034 |
|  |  | Vaporized | 2 mg | 30 min | +21.0 [-9.1, +51.0] | +0.68 | .153 |
|  |  |  | 4 mg | 30 min | +35.1 [+12.2, +58.1] | +1.49 | .007 |
|  | Stimulatory (VAS) | Oral | 5 mg | 150 min | +25.3 [+9.0, +41.5] | +1.19 | .007 |
|  |  |  | 10 mg | 120 min | +33.3 [+15.3, +51.3] | +1.41 | .002 |
|  |  | Vaporized | 2 mg | 5 min | +19.0 [-5.4, +43.4] | +0.73 | .114 |
|  |  |  | 4 mg | 5 min | +42.0 [+20.7, +63.3] | +1.76 | .001 |
|  | Aversive (VAS) | Oral | 5 mg | 60 min | +6.6 [+0.1, +13.1] | +0.68 | .046 |
|  |  |  | 10 mg | 360 min | +1.7 [-3.1, +6.5] | +0.24 | .459 |
|  |  | Vaporized | 2 mg | 5 min | +3.3 [-2.7, +9.3] | +0.50 | .234 |
|  |  |  | 4 mg | 60 min | +10.5 [+1.3, +19.7] | +0.91 | .028 |
| Reinforcement | MCP crossover (USD) | Oral | 5 mg | — | -0.4 [-7.5, +6.7] | -0.04 | .911 |
|  |  |  | 10 mg | — | +2.1 [-4.9, +9.1] | +0.21 | .533 |
|  |  | Vaporized | 2 mg | — | +4.8 [-0.1, +9.7] | +0.70 | .052 |
|  |  |  | 4 mg | — | +9.0 [+3.6, +14.5] | +1.05 | .004 |
| Cognition | HVLТ-R Total (words) | Oral | 5 mg | — | +0.9 [-3.2, +5.0] | +0.14 | .644 |
|  |  |  | 10 mg | — | -1.3 [-5.4, +2.7] | -0.22 | .489 |
|  |  | Vaporized | 2 mg | — | +2.2 [-4.4, +8.7] | +0.36 | .475 |
|  |  |  | 4 mg | — | +1.2 [-3.3, +5.8] | +0.28 | .570 |
|  | HVLТ-R Delayed (words) | Oral | 5 mg | — | +1.0 [-0.4, +2.4] | +0.33 | .139 |
|  |  |  | 10 mg | — | +0.2 [-1.2, +1.6] | +0.06 | .755 |
|  |  | Vaporized | 2 mg | — | +0.8 [-1.7, +3.2] | +0.32 | .484 |
|  |  |  | 4 mg | — | +0.1 [-1.4, +1.7] | +0.07 | .867 |
|  | CPT throughput | Oral | 5 mg | — | -2.2 [-17.6, +13.1] | -0.11 | .757 |
|  |  |  | 10 mg | — | -0.5 [-15.8, +14.8] | -0.02 | .943 |
|  |  |  | 2 mg | — | +2.7 [-9.8, +15.3] | +0.12 | .642 |
| Domain | Outcome | Route | Dose | Peak time | MD [95% CI] | Hedges' g | p |
|  |  | Vaporized | 4 mg | — | -1.1 [-12.7, +10.6] | -0.06 | .842 |
| Cardiovascular | Systolic BP<br>(mmHg) | Oral | 5 mg | 360 min | -0.8 [-10.9, +9.2] | -0.07 | .857 |
|  |  |  | 10 mg | 360 min | +0.4 [-9.5, +10.2] | +0.03 | .940 |
|  |  | Vaporized | 2 mg | 5 min | -1.1 [-8.2, +6.1] | -0.12 | .750 |
|  |  |  | 4 mg | 3 min | -8.1 [-18.7, +2.5] | -0.61 | .118 |
|  | Diastolic BP<br>(mmHg) | Oral | 5 mg | 120 min | +0.3 [-6.4, +7.0] | +0.03 | .927 |
|  |  |  | 10 mg | 240 min | +0.6 [-5.2, +6.4] | +0.08 | .831 |
|  |  | Vaporized | 2 mg | 3 min | -5.3 [-12.2, +1.7] | -0.77 | .124 |
|  |  |  | 4 mg | 5 min | -5.9 [-15.2, +3.4] | -0.59 | .192 |
|  | Heart rate<br>(BPM) | Oral | 5 mg | 5 min | +0.2 [-7.1, +7.4] | +0.02 | .956 |
|  |  |  | 10 mg | 360 min | +0.5 [-6.7, +7.7] | +0.04 | .884 |
|  |  | Vaporized | 2 mg | 15 min | +1.0 [-6.6, +8.5] | +0.12 | .780 |
|  |  |  | 4 mg | 5 min | **+11.6 [+3.7, +19.5]** | **+1.35** | **.009** |

In the vaporized THC arm, peak Pleasurable and Stimulatory ratings differed by dose (p=.019 and .004, respectively). Ratings rose within 3 to 5 min and peaked between 5 and 30 min, substantially earlier than the oral arm. At peak, Pleasurable ratings were higher than placebo by 35.1 points at 4 mg (p=.007, g=1.49) and Stimulatory ratings by 42.0 points (p=.001, g=1.76); neither differed significantly at 2 mg (p=.15 and .11). Aversive ratings exceeded placebo at 4 mg by 10.5 points (p=.028, g=0.91).

### 3.4 Reinforcement Value and Abuse Liability

Vaporized but not oral THC increased reinforcement value, measured using the MCP (**Figure S5**). The oral THC arm showed no dose effect (p=.67) and no active dose differed from placebo (all p≥.53). In the vaporized THC arm, a significant overall dose effect (p=.018) was driven by a higher crossover value after 4 mg than placebo, by about $9 (p=.004, g=1.05).

### 3.5 Cognitive Performance

Neither oral nor vaporized THC was associated with changes in sustained attention (CPT throughput), verbal learning (HVLT-R Total Recall), or memory (HVLT-R Delayed Recall) (all dose effects p≥.21; **Figure S6**). No active dose differed from placebo on any cognitive measure.

### 3.6 Heart Rate and Blood Pressure

The route of administration of THC produced distinct effects on HR and BP (**Figure 2D-F**). In the oral THC arm, peak HR, SBP, and DBP responses did not differ by dose (p=.99, .96, and .98, respectively).

In the vaporized THC arm, peak HR response differed by dose (p=.006). At 4 mg, the peak HR rise exceeded placebo by 11.6 bpm (p=.009, g=1.35), but not at 2 mg (p=.78). Peak SBP and DBP responses did not differ by dose (p=.28 and .26).

### 3.7 Exposure–Response

Higher peak active combined analytes concentration (Cmax) was associated with greater Pleasurable and Stimulatory effects after both routes, and these associations were numerically stronger after vaporization than after oral dosing (Pleasurable oral p=.019, vaporized p<.001; Stimulatory oral p=.001, vaporized p<.001). HR was strongly associated with exposure after vaporization (p=.001) but not after oral dosing, and neither BP measure tracked exposure after either route (**Section S16**, **Table S8**).

### 3.8 Adverse Events and Blinding

Five treatment emergent AEs occurred across 58 sessions, all at vaporized 4 mg and concentrated on two participants (**Table S4**). One participant experienced four AEs: bradycardia (HR <60 bpm), drowsiness, dizziness/lightheadedness, and a vasovagal syncope that led to study withdrawal after the first session. A second participant experienced mild dizziness/lightheadedness. No AEs occurred in the remaining 56 test sessions.

Blinding was generally maintained, except for vaporized 4 mg, which was correctly identified by 80% of participants and raters (BI=0.6; **Table S5**).

## 4. Discussion

In this randomized, double-blind, placebo-controlled trial, we found that route of administration determined both the magnitude of THC exposure and metabolite formation in adults aged ≥65 years, and that those differences carried through to abuse liability and cardiovascular response, but not to cognitive performance. Collectively, these results extend a sparse evidence base on the effects of cannabinoids among older adults, among whom only oral and oromucosal formulations had been studied [9,38–40,71]. By characterizing the effects of oral and vaporized THC across pharmacokinetics, subjective effects, reinforcement value, cognitive performance, HR and BP, and tolerability, we demonstrate that oral and vaporized THC should not be treated as interchangeable when assessing risks among older adults. These data also provide a benchmark for future studies of THC- and cannabis-based products in this population.

### 4.1 Pharmacokinetic Effects of Route of Administration

For oral THC, our observed Cmax and Tmax match dronabinol pharmacokinetics reported in healthy adults aged 18-55 at the same 5 mg dose (Cmax 2.20-2.61 ng/mL, Tmax 1.0-1.5 h)[72]. Our observed Cmax sits at the upper end of the range reported in the only prior controlled oral THC study in adults ≥65 (oromucosal Namisol)[38], though differing bioavailability limits dose-by-dose comparison. Oral dosing also generated 11-OH-THC at peak concentrations comparable to THC and total exposure approximately 1.5-fold higher (**Figure 1**), reflecting extensive first-pass metabolism. Oral THC exposure was lower in participants with higher body weight (**Section S17**), consistent with a larger volume of distribution at a fixed dose.

Vaporized THC produced rapid-onset, THC-dominant exposure (Cmax 24.6 and 53.8 ng/mL at 2 and 4 mg; Tmax 3 min), with minimal 11-OH-THC. Because 11-OH-THC is a CB1-receptor agonist at least as active as THC[73,74], the two routes differed not only in total exposure but also in the relative contribution of CB1-active cannabinoids. Prior vaporized-THC studies in younger adults used cumulative within-session dosing[75], precluding characterization of the concentration–time profile of an individual dose; our single-dose design isolates that profile. Other studies used whole-plant cannabis in younger cohorts[49,76], often as whole-blood analytes, roughly half plasma values[77].

### 4.2 Abuse Liability Across Routes of Administration

Abuse liability, reflected in subjective effects and reinforcement value, tracked each route’s pharmacokinetics, consistent with adult laboratory studies of oral THC[14] and of oral and vaporized cannabis[50]. The magnitude of subjective effects also scaled with exposure: pleasurable and stimulatory effects, but not aversive effects, increased with peak active cannabinoid concentration (Cmax) after both routes, a relationship that changed little whether 11-OH-THC was excluded or weighted up to twice THC (**Section S16).**

Because faster delivery and more rapid onset are associated with greater acute reinforcement[82], vaporization strengthens the temporal coupling between administration and effect. Consistent with this, prior work found only modest abuse liability for oral dronabinol at 10 and 20 mg in adults with daily cannabis use[16], and in the current study vaporized THC 4 mg increased the hypothetical monetary value assigned to the drug experience—whereas neither oral dose did.

### 4.3 Cognitive Performance in the Context of Aging

Among our sample of healthy older adults, neither oral nor vaporized THC impaired sustained attention, verbal learning, or memory. The evidence base in older adults remains sparse[83]. Impairment is clearer in younger samples given higher doses: oral THC at 15 mg impaired episodic memory and learning[84]. Vaporized cannabis at 20 and 40 mg impaired multiple verbal memory domains[85]. Most directly, adults aged 55-70 showed less acute impairment in learning and processing speed than adults aged 21-25 at equivalent plasma THC concentrations[20].

Age-related declines in CB1 receptor expression provide a plausible substrate for these differences in acute THC effects across the lifespan[18,19]. This may be especially relevant for episodic memory: the hippocampus, which supports the verbal learning probed by the HVLT-R, is among the brain’s richest CB1-receptor regions[86], and verbal memory impairment is among the most consistent acute effects of THC in younger adults[13,87]. Reduced hippocampal CB1 binding could therefore blunt this effect, consistent with what we observed. Additionally, chronic low dose THC reversed age-related cognitive decline in aged but not young animals[88,89].

### 4.4 Heart Rate, Blood Pressure, and Adverse Events

The absence of HR elevations after oral THC in our study contrasts with younger adult findings, among whom oral THC at 7.5 and 15 mg dose-dependently increased HR peaking 90-180 min after dosing[14]. In contrast, the observed vaporized pattern is consistent with the rapid-onset tachycardia reported after inhaled cannabis in adult laboratory studies[49].

In the exposure–response analysis, HR increased with peak THC exposure after vaporization, whereas neither BP measure tracked exposure after either route. This concentration-dependence aligns with concentration-effect modeling of intravenous THC in healthy volunteers, in which HR rose alongside rising THC and 11-OH-THC plasma levels[90]. Cannabis-associated tachycardia is autonomically mediated and has been attributed to sympathetic activation and parasympathetic withdrawal[91]. CB1 receptor signaling is implicated in this autonomic cardiovascular control[92]. Because cardiovascular reserve and autonomic regulation decline with age, the rapid HR increase following vaporized THC may have greater clinical relevance in older adults at risk for orthostatic symptoms and falls[25].

All five AEs observed across 58 sessions occurred at vaporized THC 4 mg, in two participants. One was a syncopal episode, the only severe event (**Table S4**). The other events were consistent with the dizziness and nausea profile reported in real-world medical cannabis use among older adults[93]. Their concentration at vaporized THC 4 mg under controlled conditions cautions against higher inhaled doses in this age group.

### 4.5 Clinical and Research Implications for Older Adults

The primary practical implication of the present study is that oral and vaporized THC should not be treated as interchangeable in risk assessment for older adults. Compared with oral dosing, vaporization produced approximately 9-fold higher peak THC concentrations and subjective effects that peaked approximately 2 hours earlier. At 4 mg, vaporized THC also increased reinforcement value (+$9.0) and HR (+11.6 bpm); effects not observed after oral THC.

When the medical goal is gradual symptom relief with minimal acute cardiovascular and abuse-liability concerns, oral formulations may offer a more predictable risk profile, although their slow onset may prompt a second dose before the first takes effect. Vaporized products require caution because even low milligram doses can produce rapid peak exposure, stronger temporal coupling between administration and effect, and acute cardiovascular changes. Thus, dose selection should account for route, rate of delivery, achieved peak exposure, and patient-level vulnerability to dizziness, orthostatic symptoms, falls, or cardiovascular events.

### 4.6 Limitations and Future Directions

Some limitations warrant discussion. Regarding the sample, participants were required to have prior cannabis exposure, although cannabis use disorder was excluded and participants tested negative before each session. Findings may therefore not generalize to cannabis-naive older adults, in whom acute subjective responses may differ: among cannabis-naive older adults given oral Namisol at comparable plasma Cmax, only 4 of 11 endorsed any “feeling high”[38]; the same investigators reported significant responses in cannabis-experienced young adults[78]. Prior familiarity could influence how THC effects are recognized or reported[79–81], but this could not be distinguished from THC’s pharmacologic effects here. Findings may also not extend to older adults with cardiovascular or hepatic conditions, or those taking medications that interact with THC metabolism. Regarding design, the acute single-dose protocol did not address repeated dosing, steady-state pharmacokinetics, or cumulative cardiovascular risk; and because route was a between-participant factor, cross-route contrasts should be interpreted accordingly. Regarding assessment, the MCP captured hypothetical rather than actual drug choice, and cognition was assessed at a single post-dose timepoint.

Future studies should examine steady-state pharmacokinetics after repeated dosing and cumulative cardiovascular and cognitive effects. A younger comparison arm would allow age-specific effects to be tested directly rather than inferred from cross-study comparison, and a fully within-subject crossover would sharpen cross-route comparison, and larger, more diverse samples are needed, including clinical populations with chronic pain, sleep disturbance, or dementia-related behavioral symptoms.

### 4.7 Conclusions

Among healthy adults aged ≥65 years, oral and vaporized THC produced distinct pharmacokinetic and pharmacodynamic profiles, differing most in onset, metabolite formation, abuse liability, and acute cardiovascular effects.

Oral THC produced delayed, prolonged exposure with substantial 11-OH-THC formation and slow-rising subjective effects, without increases in reinforcement value, HR, or adverse events; vaporized THC produced rapid, THC-dominant exposure, rapid-onset subjective effects, higher reinforcement value at 4 mg, acute HR elevation, and all observed adverse events.

Despite demonstrable exposure, our sample did not experience cognitive impairment after either route.

Together, these findings provide controlled reference data for THC pharmacokinetics and pharmacodynamics in adults aged ≥65 years and indicate that route of administration should be considered an independent determinant of THC exposure and clinical risk in older adults, rather than simply an alternative method of delivering equivalent doses. Extending this benchmark to repeated dosing, clinical populations, and formulations beyond purified THC is the next step toward evidence-informed cannabinoid use in older adults.

## Data Availability Statement

The data that support the findings of this study are available from the corresponding author upon reasonable request.

## Acknowledgments

We thank Christina Riggione, Jocelyn Ra, Enoch Ofori, Mayte A. Cerezo-Matias, and Becky Suh for their contributions to data collection. We are grateful to Susan Gray, BSN, RN, Angelina Genovese, RNC, BSN, MBA, and Vanessa Hurley, RN, for nursing support. We also thank the participants and the staff of the Biological Studies Unit at the VA Connecticut Healthcare System.

## Author Contributions

G.P.A.C. contributed to data analysis, and writing of the original manuscript. S.A., J.M., S.D., S.M., and J.N. contributed to data collection and study conduct. R.G. supervised the statistical analyses. M.M.R. contributed to the pharmacokinetic analyses. T.E. and H.N. contributed to the bioanalytical analyses. M.S. provided supervision and critical revision of the manuscript. J.P.D. was responsible for the conceptualization, supervision, funding acquisition, project administration, and writing, reviewing, and editing of the final manuscript. All authors gave final approval for the version to be published and agree to be accountable for all aspects of the work.

## Funding

This work was supported by grant R21DA057240 from the National Institute on Drug Abuse (NIDA) to Dr. De Aquino. The other authors received no specific funding for this work.

## Competing Interests

Dr. De Aquino has received research support from Jazz Pharmaceuticals and Ananda Scientific and has served as a paid consultant for Boehringer Ingelheim. The remaining authors declare no conflict of interest.

## Notes

### Clinical Trial

NCT05906511

### Clinical Protocols

https://clinicaltrials.gov/study/NCT05906511

### Author Declarations

The study protocol was approved by the Institutional Review Board of Yale University, New Haven, Connecticut, United States, and by the Institutional Review Board of the VA Connecticut Healthcare System, West Haven, Connecticut, United States. Ethical approval was granted by both boards. All participants provided written informed consent before any study procedures were performed.

