## Supplement for "Pharmacokinetics and Pharmacodynamics of Oral and Vaporized Δ9-Tetrahydrocannabinol in Older Adults"

Supplementary Material

Pharmacokinetics, Subjective Effects, and Abuse Liability of Oral and Vaporized Δ9-Tetrahydrocannabinol in Adults Aged 65 Years and Older

**S1. Inclusion and Exclusion Criteria**

**Inclusion criteria:**

1. Healthy adults aged ≥65 years.
2. Prior exposure to THC or cannabis: at least once in the last 10 years; 1-10 times in the last 20 years; or more than 20 times lifetime.
3. Capable of providing written informed consent in English.

**Exclusion criteria:**

1. Meeting DSM-5 criteria for psychiatric or substance use disorders (other than tobacco use disorder) within the last year.
2. Current cannabinoid use, evidenced by urine drug screen.
3. Having a history of treatment for cannabis use disorder, or, history of intent or current intent of abstaining from cannabis use.
4. Clinically significant medical disorders (e.g., liver or kidney dysfunction, COPD or asthma, immunosuppressing conditions, history or presence of epilepsy, seizures, or head trauma with loss of consciousness).
5. Neurological conditions that may change the response to nociceptive stimuli (e.g., stroke, peripheral neuropathy) or that lead to loss of balance, evidenced by a neuro-sensory exam.
6. Contraindications for exposure to nociceptive stimuli, such as untreated hypertension.
7. Current regular use of drugs known to affect pain or that are prominent inducers or inhibitors of CYP2C9, CYP3A4, or UGT1A9 (e.g., carbamazepine, valproate, fluvoxamine, paroxetine).
8. Major neurocognitive disorders precluding participation, evidenced by clinical exam.
9. Abnormal EKG, arrythmia, vasospastic disease, chronic heart failure, or presence of a pacemaker.
10. Personal or family history of primary psychotic disorders, or mood disorders with psychotic features.
11. Current suicidal ideation.
12. Allergy or serious adverse reactions to sesame oil, THC, or cannabis.
13. Receipt of any drug as part of a research study within 30 days before study medication administration.
14. Medical conditions that increase the risk of respiratory problems (e.g. COPD, asthma, recuring bronchitis, reactive airway disorder). (Applies only to vaporized arm)
15. History of environmental sensitivities (e.g. bronchospastic allergies, multiple chemical sensitivities) or other airway sensitivities that require the use of an epi pen. (Applies only to vaporized arm)

**S2. Drug Preparation Procedures**

**Oral arm (dronabinol).** Oral THC was administered as dronabinol, an FDA-approved Schedule III synthetic Δ9-tetrahydrocannabinol capsule formulation (Marinol® or generic equivalent; distributor McKesson Corporation, Irving, TX, USA). Bioavailability is approximately 20-25%; median Tmax is approximately 2.5 hours in the fasted state and approximately 5.6 hours fed; elimination half-life is 25-36 hours; hepatic metabolism involves CYP3A4, CYP2C9, and UGT1A9. Doses of 5 mg and 10 mg were prepared as over-encapsulated capsules visually identical to matching placebo capsules. Capsules were prepared and dispensed by the research pharmacy, which exclusively held the randomization sequence. On each test day, the study nurse or physician administered the capsule after baseline assessments in the fasted state; participants received a standardized meal after the 4-hour timepoint. Conditions were delivered in counterbalanced random order across three test days separated by at least 72 hours.

**Vaporized arm (purified Δ9-THC).** Vaporized THC was delivered using the Volcano Hybrid® vaporizer (Storz & Bickel, Tuttlingen, Germany), a device validated for reproducible, smoke-free cannabinoid delivery. Pharmaceutical-grade Δ9-tetrahydrocannabinol (99% purity) was compounded as a 10 mg/mL ethanolic solution by Hybrid Pharma LLC (Deerfield Beach, FL, USA) under DEA Schedule I license and dispensed in visually identical syringes by the research pharmacy. The vaporizer was preheated to 226 ± 5 °C; the prescribed dose of ethanolic THC solution was loaded into the vaporization chamber and vaporized into the Volcano filling-chamber balloon. Participants then inhaled the balloon contents using a standardized cued-puff procedure: a 5-second inhalation, 10-second breath-hold, and 45-second exhalation/wait, with the 60-second cycle repeated until the balloon was emptied. Active doses of 2 mg and 4 mg were selected to approximate the systemic exposure of the oral 5 mg and 10 mg doses and to fall within the low-to-moderate range characterized in prior human laboratory studies of inhaled THC. Placebo sessions used a matched ethanolic vehicle (no THC) delivered through the same Volcano device with the same cued-puff protocol.

**Allocation concealment.** Conditions were delivered in counterbalanced random order across three test days per arm, separated by at least 72 hours. Participants, raters, and the principal investigator were blinded to condition on each test day; the randomization sequence was held exclusively by a research pharmacist. End-of-session blinding assessment (Table S5) confirmed concealment for oral active doses and vaporized placebo and 2 mg.

**S3. Assessment Schedule**

**Table S1.** Per-assessment timepoint schedule by arm. Filled cell indicates the assessment was administered at that timepoint; empty indicates not administered. PK = plasma pharmacokinetic sample; DEQ = Drug Effects Questionnaire; VS = vital signs (HR, SBP, DBP).

| Session Assessment Schedule by Arm | | | | | |
| --- | --- | --- | --- | --- | --- |
| Time | Oral PK | Oral DEQ | Vap PK | Vap DEQ | VS (both arms) |
| Pre-dose baseline | ● | ● | ● | ● | ● |
| 0 (dose) |  |  |  |  | ● |
| 3 min |  | ● | ● | ● | ● |
| 5 min |  | ● | ● | ● | ● |
| 15 min |  | ● | ● |  | ● |
| 30 min |  | ● | ● | ● | ● |
| 45 min | ● | ● | ● | ● | ● |
| 60 min | ● | ● | ● | ● | ● |
| 90 min | ● | ● |  | ● | ● |
| 120 min | ● | ● |  | ● | ● |
| 150 min | ● | ● |  | ● | ● |
| 180 min | ● | ● |  | ● | ● |
| 210 min | ● | ● | ● | ● | ● |
| 240 min | ● | ● | ● | ● | ● |
| 360 min | ● | ● | ● | ● | ● |
| 480 min | ● | ● | ● | ● | ● |
| 1440 min (24-h) | ● |  | ● |  | ● |
| **Total per session** | **12** | **15** | **12** | **14** | **17** |

When intravenous access could not be obtained for plasma sampling, the affected session used individual venipunctures at a subset of the scheduled timepoints; this occurred once in the study (vaporized 2 mg session).

**S4. Cognitive Battery Implementation Details**

**Continuous Performance Test (CPT, ANAM).** Sustained attention was assessed with the Continuous Performance Test from the Automated Neuropsychological Assessment Metrics (ANAM) battery. Participants viewed a sequential stream of stimuli and responded to a designated target via key press. Throughput (correct responses per minute, combining accuracy and reaction time) served as the primary outcome.

**Hopkins Verbal Learning Test - Revised (HVLT-R).** Verbal learning and memory were assessed with the HVLT-R. Participants heard a 12-word list across three immediate-recall trials; Total Recall was the sum across the three trials (range 0-36). A delayed-recall trial after approximately 20 minutes yielded Delayed Recall (range 0-12). Alternate forms were administered in counterbalanced order across the three test sessions per arm to minimize practice effects.

**S5. Participant Flow**

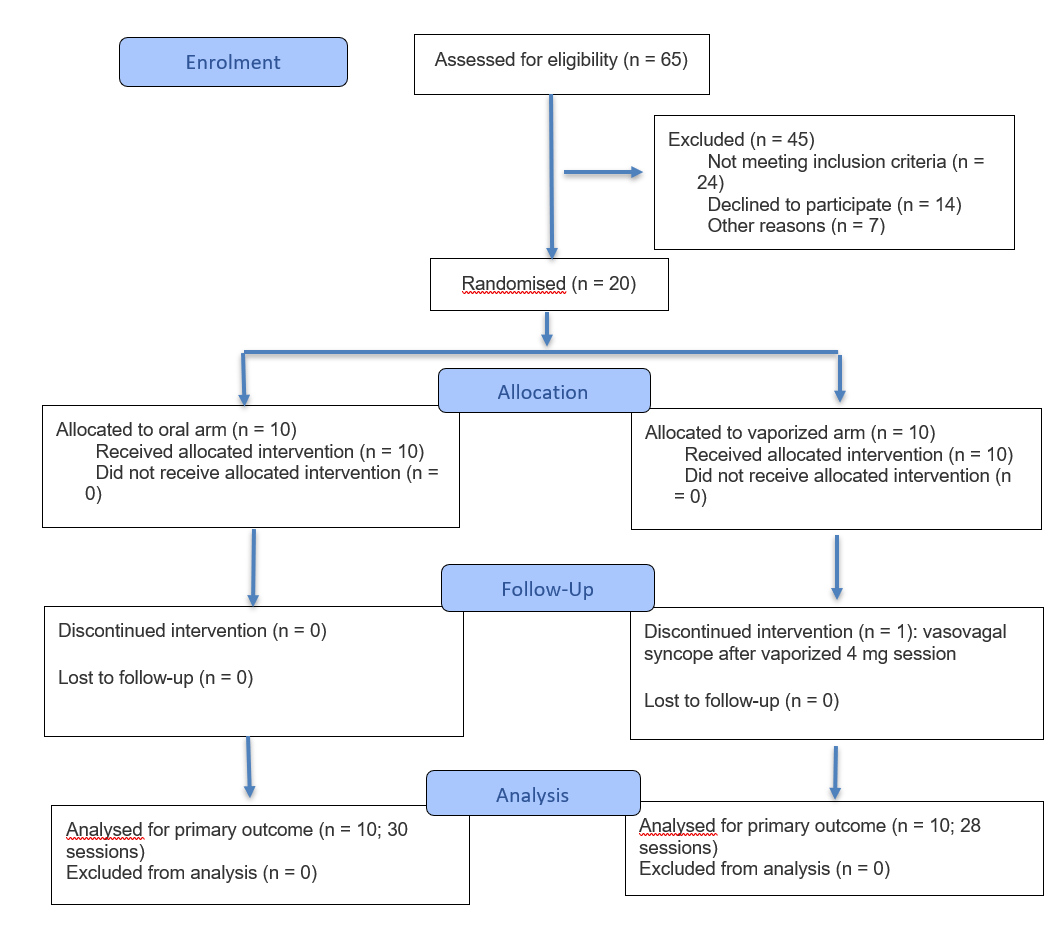
**Figure S1.** **CONSORT 2025 Flow Diagram.** Flow diagram of the progress through the phases of a randomised trial of two groups (that is, enrolment, intervention allocation, follow-up, and data analysis).

**S6. Plasma Pharmacokinetics — Bioanalytical Assay and Non-Compartmental Analysis**

**Non-compartmental analysis.** Plasma THC, 11-OH-THC, and THC-COOH were extracted by solid-phase extraction (Oasis PRiME HLB µElution plate; Waters Corporation, Milford, MA) and separated on an ACQUITY UPLC I-Class system with an ACQUITY UPLC BEH C18 column (2.1 × 50 mm, 1.7 µm; Waters Corporation) using a gradient of water and acetonitrile, each containing 0.1% formic acid. Detection used a Xevo TQ-XS triple-quadrupole mass spectrometer (Waters Corporation) in positive-mode electrospray ionization, with quantification by multiple reaction monitoring (THC, m/z 315.1 > 193.2; 11-OH-THC, m/z 331.3 > 313.1; THC-COOH, m/z 345.3 > 327.3) against deuterated internal standards (THC-d3, 11-OH-THC-d3, and THC-COOH-d3). Lower limits of quantification were 0.2, 1.0, and 0.5 ng/mL for THC, 11-OH-THC, and THC-COOH, respectively. Non-compartmental analysis was performed using the *PKNCA* package v0.12.1 in R version 4.5. The linear-trapezoidal method was applied with start-of-interval concentration imputed to zero when not observed.

**S7. Plasma Pharmacokinetics - 24-Hour**

Each test session was followed by a 24-hour (1440-minute) follow-up visit at which plasma was sampled. The 24-hour data are not part of the primary 0-480 min on-dose analyses; this section provides descriptive values for completeness (Table S3).

**Table S2.** Mean ± SD plasma concentrations at the 24-hour follow-up visit, by arm, dose, and analyte.

| 24-Hour Plasma Concentrations by Route and Dose | | | |
| --- | --- | --- | --- |
| Analyte | Arm | Dose | C24h mean ± SD (ng/mL) |
| **Δ9-THC** | Oral | Placebo | 0.13 ± 0.16 |
|  | Oral | 5 mg | 0.18 ± 0.11 |
|  | Oral | 10 mg | 0.33 ± 0.54 |
|  | Vaporized | Placebo | 0.28 ± 0.32 |
|  | Vaporized | 2 mg | 0.31 ± 0.35 |
|  | Vaporized | 4 mg | 0.72 ± 0.59 |
| **11-OH-THC** | Oral | Placebo | 0.00 ± 0.00 |
|  | Oral | 5 mg | 0.50 ± 0.00 |
|  | Oral | 10 mg | 0.98 ± 1.05 |
|  | Vaporized | Placebo | 0.00 ± 0.00 |
|  | Vaporized | 2 mg | 0.08 ± 0.20 |
|  | Vaporized | 4 mg | 0.50 ± 0.00 |
| **THC-COOH** | Oral | Placebo | 1.14 ± 1.51 |
|  | Oral | 5 mg | 3.88 ± 1.57 |
|  | Oral | 10 mg | 22.64 ± 38.53 |
|  | Vaporized | Placebo | 0.73 ± 1.06 |
|  | Vaporized | 2 mg | 0.63 ± 0.49 |
|  | Vaporized | 4 mg | 2.77 ± 1.78 |

*Parent Δ9-THC and 11-OH-THC return to or near LOQ by 24 hours after both routes. THC-COOH, the inactive carboxylic-acid metabolite, remains detectable at 24 hours after oral 10 mg (mean 22.6 ng/mL), consistent with the slow elimination of this metabolite following oral administration that generates substantial first-pass formation. Vaporized 24-hour THC-COOH values are an order of magnitude lower, consistent with the minimal first-pass formation characteristic of inhaled THC.*

**Figure S2. Individual plasma concentration**–**-time profiles of THC, 11-OH-THC, and THC-COOH by route and dose.** Thin translucent lines = individual-participant profiles; bold lines = group means (linear concentration scale). Companion to the group-mean display in main-text Figure 1.

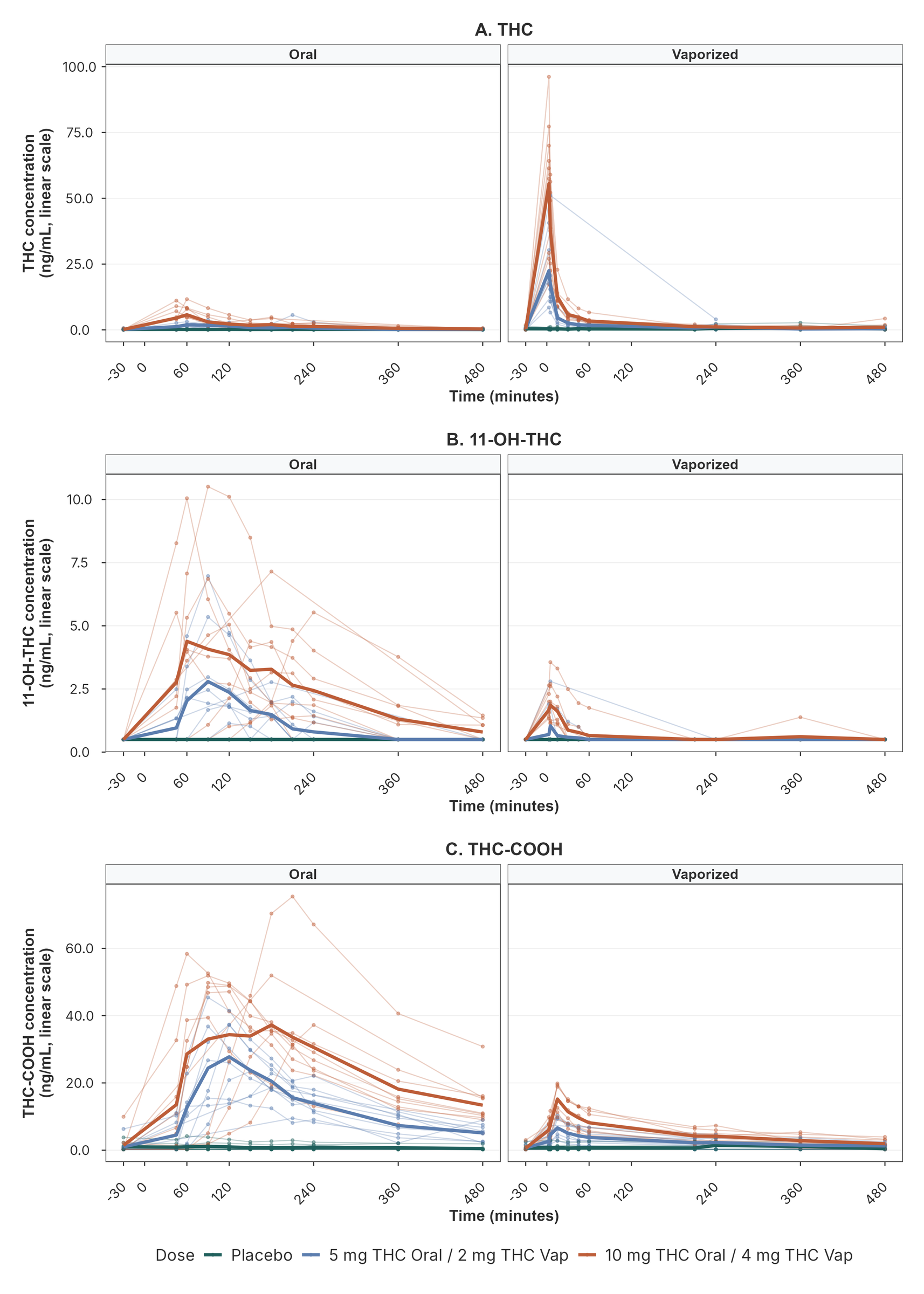

**S8. Subjective Drug Effects - Peak Change from Baseline**

**Figure S3. Per-subject** peak change from baseline in DEQ composites by dose condition (oral and vaporized arms). Peak change from baseline = the single timepoint with the largest absolute deviation from the pre-dose baseline, signed. Panels: (A) Stimulatory; (B) Pleasurable; (C) Aversive. Bars = group mean ± SE; individual participants overlaid as dots.

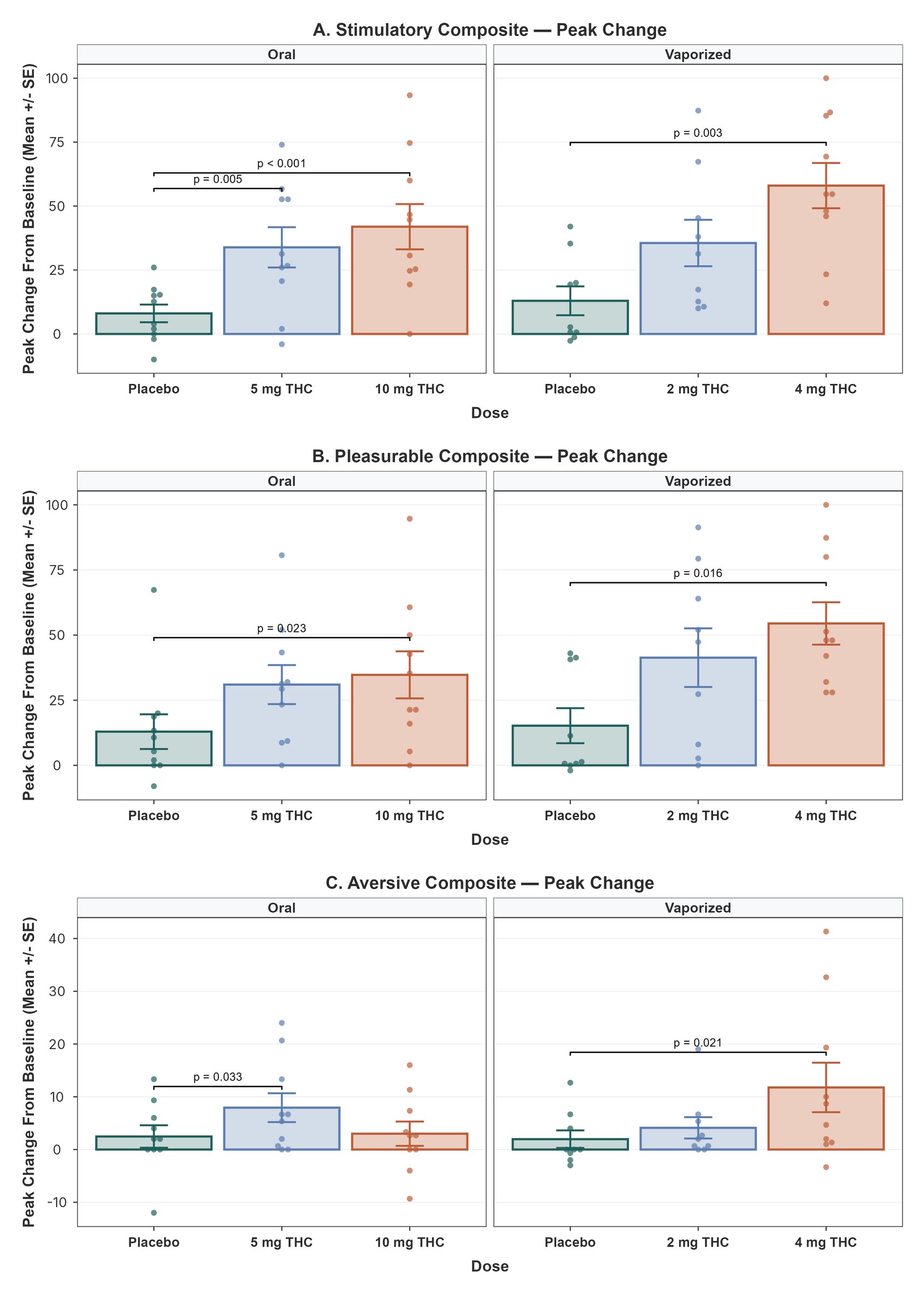

**S9. Subjective Drug Effects - Timeline with Per-Subject Trajectories**

**Figure S4. Time course of DEQ** subjective drug-effect composites with individual-participant trajectories. Mean change from baseline on the three DEQ composites: (A) Stimulatory; (B) Pleasurable; (C) Aversive. DEQ items were rated on 0-100 mm visual analog scales. Results are shown separately by route of administration: oral THC in the left panels and vaporized THC in the right panels. Thin translucent lines = individual-participant trajectories; bold lines = group means.

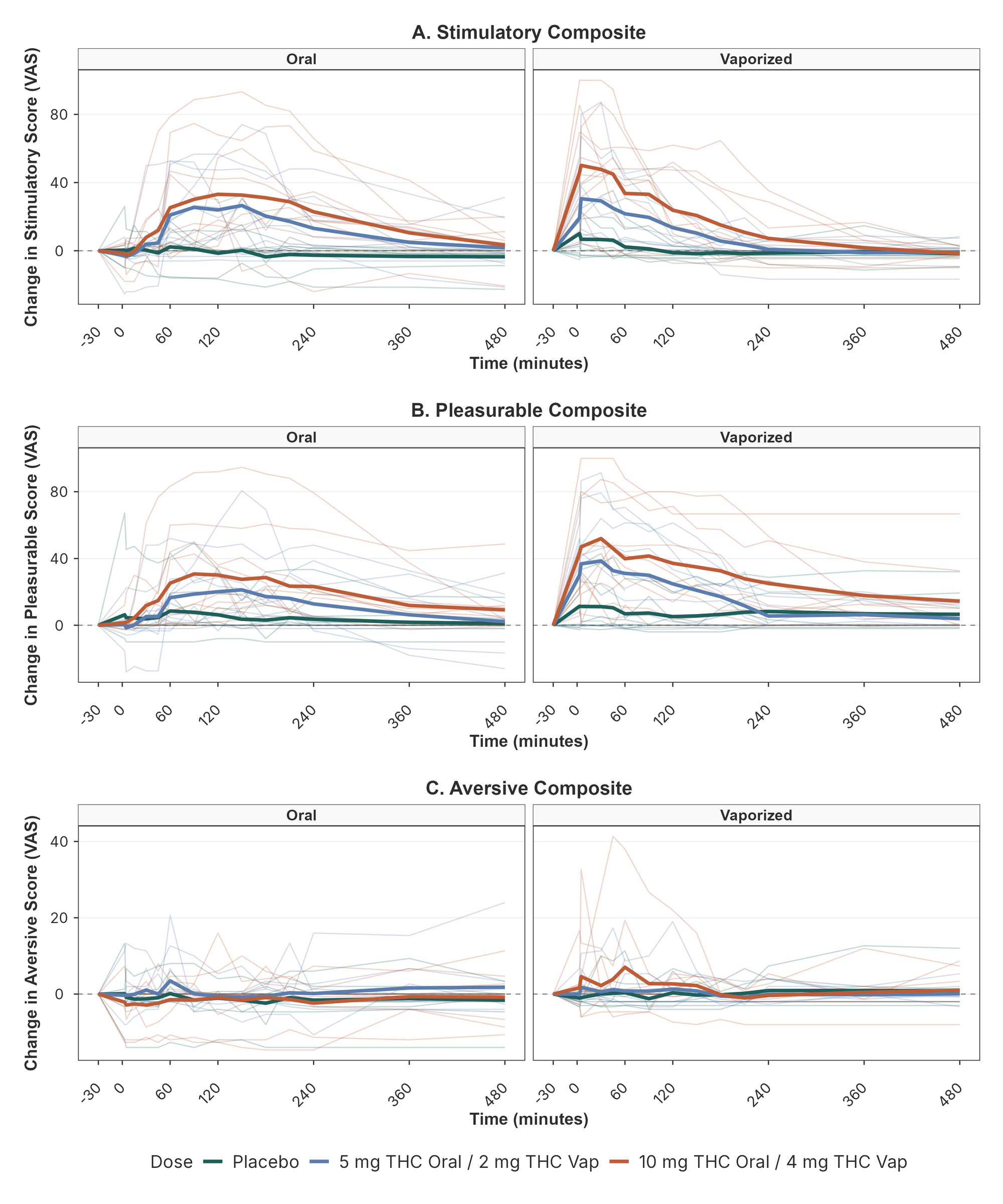

**S10. Reinforcement Value**

**Figure S5. Multiple-Choice Procedure** (MCP) crossover-point values by dose for the oral and vaporized arms. Bars = group mean ± SE; individual participants overlaid as dots. Significance brackets denote pairwise contrasts (p < 0.05) by linear mixed-effects model.

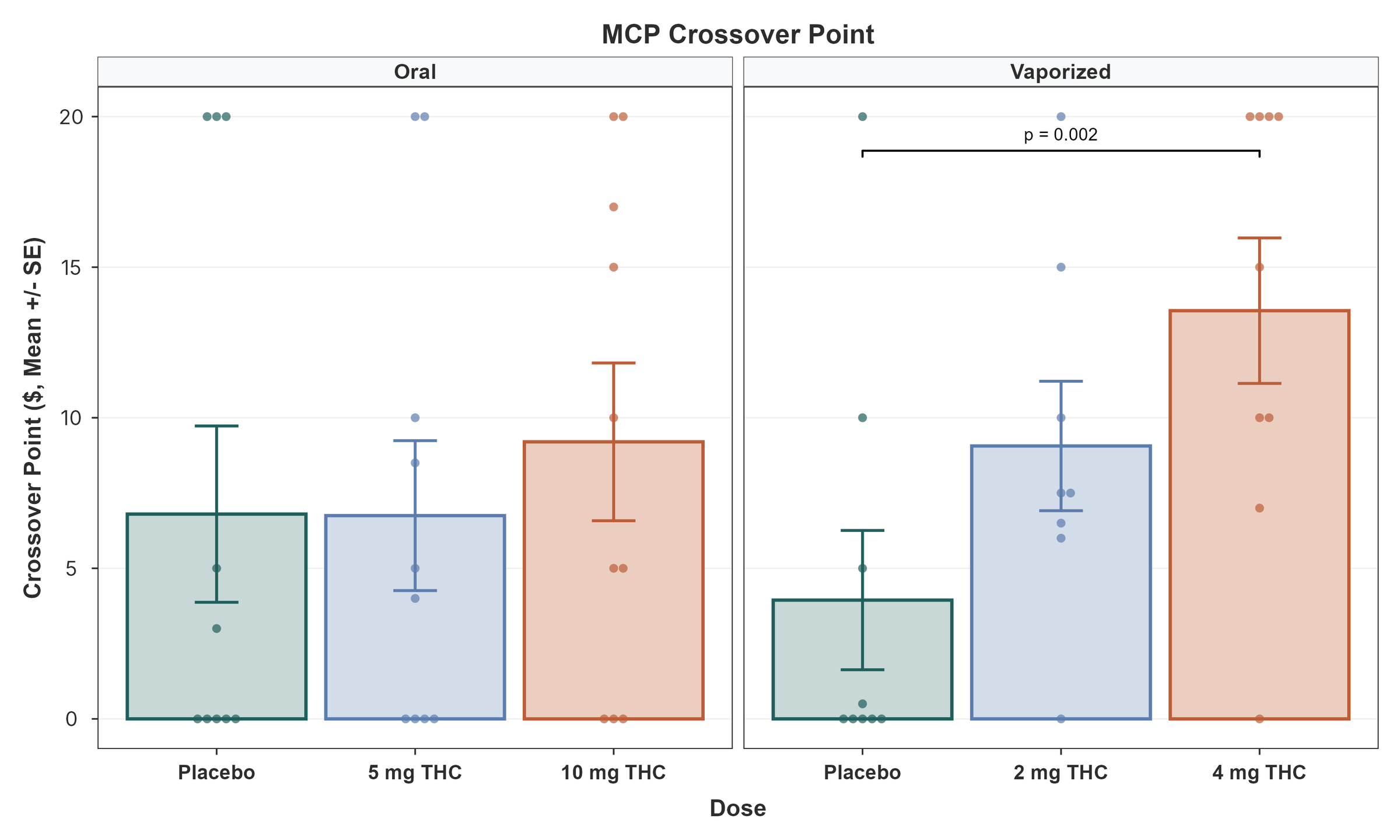

**S11. Cognitive Performance**

**Figure S6. Cognitive Performance**

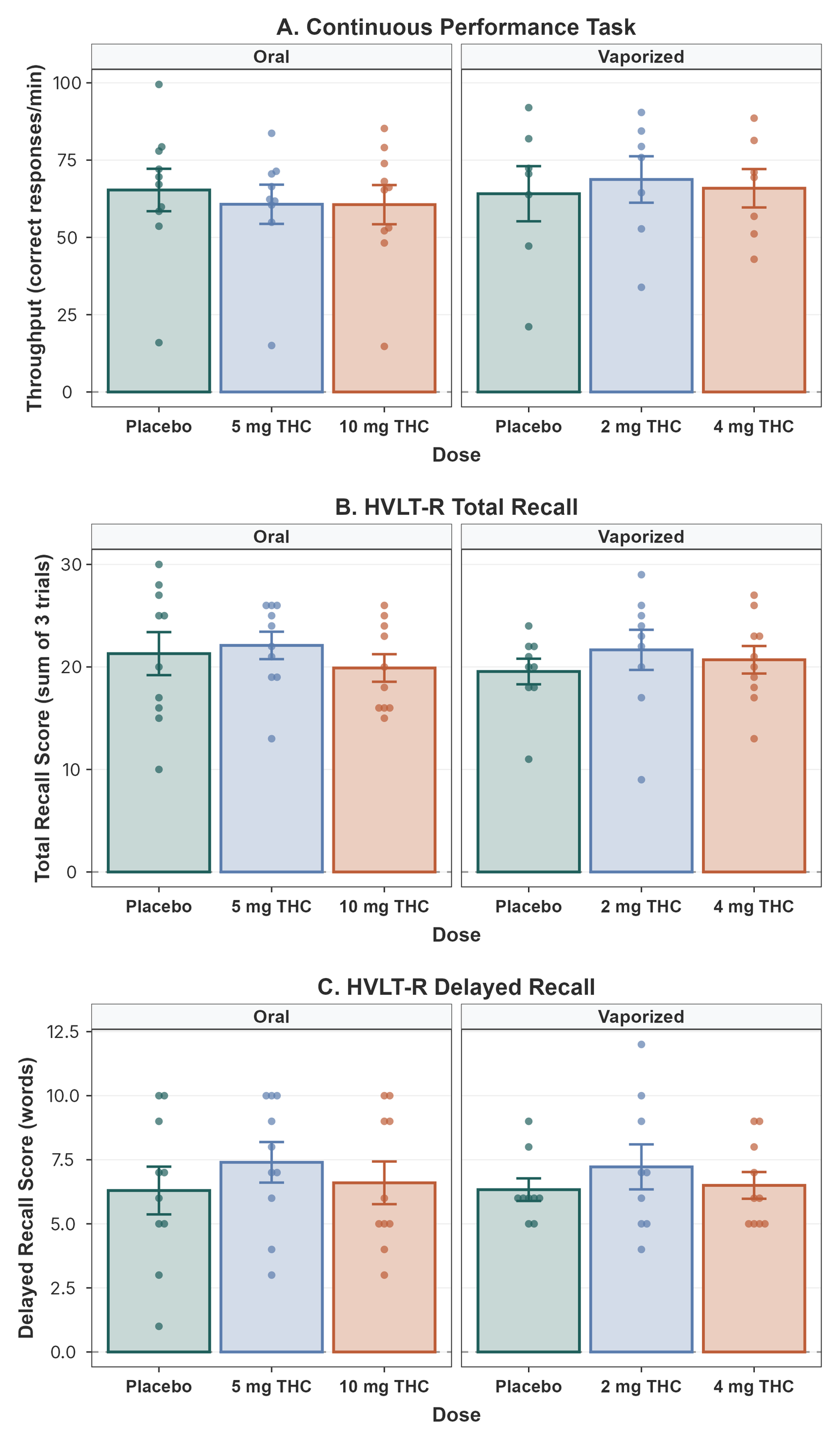

**Table S3.** Cognitive omnibus Dose effects by arm and outcome (HVLT-R Total Recall, HVLT-R Delayed Recall, ANAM CPT Throughput).

| Cognitive Omnibus Dose Effects | | | |
| --- | --- | --- | --- |
| Measure | Arm | F | p |
| **ANAM CPT Throughput** | Oral | 0.06 | .946 |
|  | Vaporized | 0.25 | .785 |
| **HVLT-R Delayed Recall** | Oral | 1.73 | .215 |
|  | Vaporized | 0.27 | .772 |
| **HVLT-R Total Recall** | Oral | 1.28 | .312 |
|  | Vaporized | 0.30 | .745 |

**S12. Heart Rate and Blood Pressure - Timeline with Per-Subject Trajectories**

**Figure S7.** Change from baseline cardiovascular time courses with individual-participant trajectories, complementing the group-mean display in main-text Figure 5. Panels: (A) Heart rate (bpm); (B) Systolic blood pressure (mmHg); (C) Diastolic blood pressure (mmHg). Thin translucent lines = individual-participant trajectories; bold lines = group means.

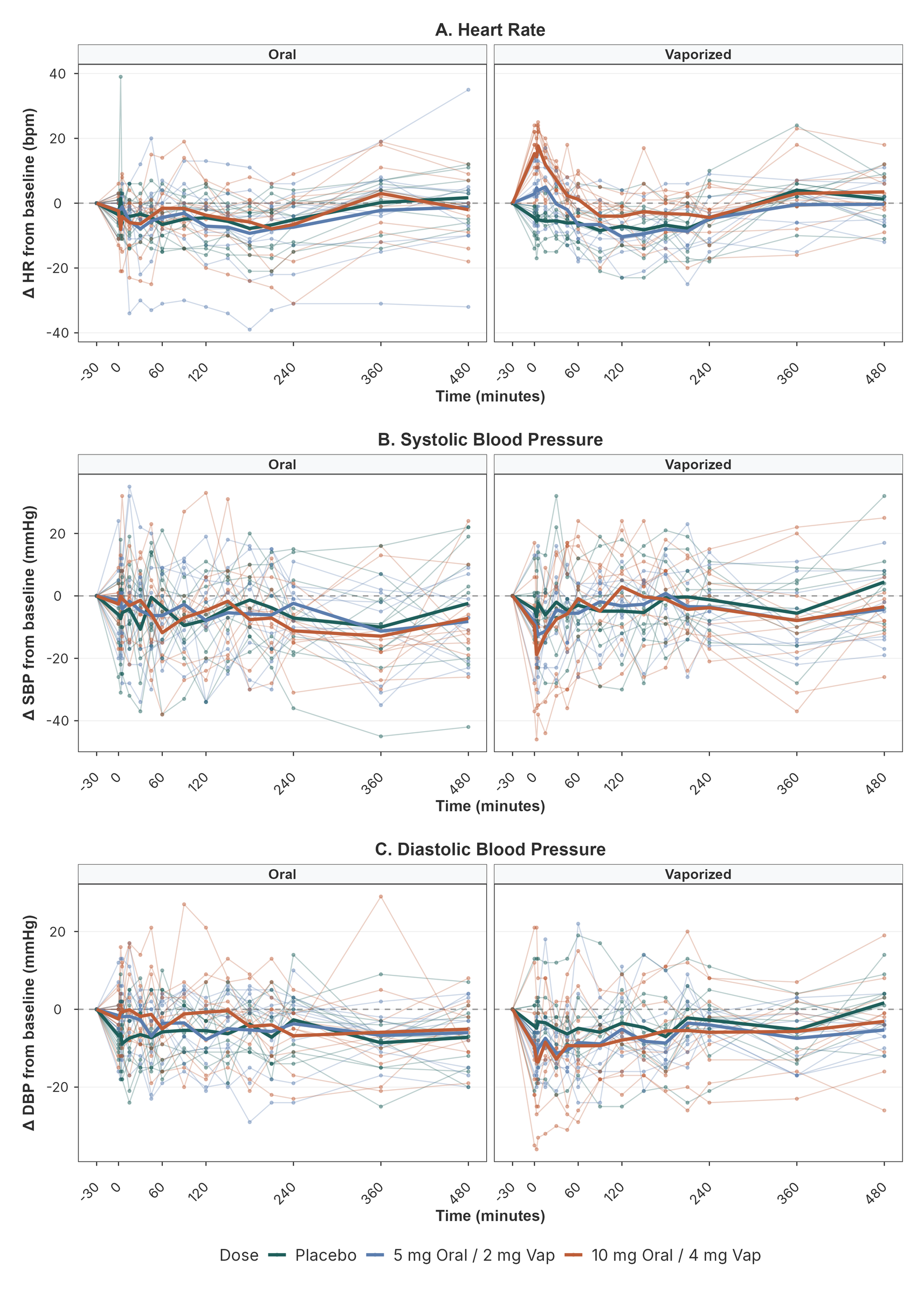

**S13. Adverse Events (SAFTEE)**

**Table S4.** SAFTEE adverse-event summary by Arm x Dose condition. N sessions = number of on-dose participant-sessions assessed; Sessions w/ any AE = sessions on which at least one item worsened from baseline; Total AE burden = summed SAFTEE severity scores across sessions; Actions taken = protocol-mandated dose holds or session pauses.

| Adverse-Event Summary by Route and Dose (SAFTEE) | | | | | |
| --- | --- | --- | --- | --- | --- |
| Arm | Dose | N sessions | Sessions w/ any AE | Total AE burden | Actions taken |
| **Oral** | **Placebo** | 10 | 0 | 0 | 0 |
| **Oral** | **5 mg THC** | 10 | 0 | 0 | 0 |
| **Oral** | **10 mg THC** | 10 | 0 | 0 | 0 |
| **Vaporized** | **Placebo** | 9 | 0 | 0 | 0 |
| **Vaporized** | **2 mg THC** | 9 | 0 | 0 | 0 |
| **Vaporized** | **4 mg THC** | 10 | 2 | 9 | 4 |

During the vaporized 4 mg test session, a participant presented a vasovagal syncope, which prompted study withdrawal.

**S14. Treatment Allocation Concealment**

**Table S5.** Treatment allocation concealment per arm x dose x role. N = number of sessions with a guess recorded; Correct (n) = sessions where the guess matched the actual mg administered; Correct (%) = same as a percentage. Cells with 50% correct (oral 5 mg and 10 mg subjects) reflect chance-level guessing; cells reaching 80% correct (vaporized 4 mg subjects and raters; oral placebo raters) reflect identifiable conditions.

| Treatment Allocation Concealment | | | | | | | | |
| --- | --- | --- | --- | --- | --- | --- | --- | --- |
|  | | | Subject guess | | | Rater guess | | |
| Arm | Dose | N | Correct (n) | Correct (%) | BI | Correct (n) | Correct (%) | BI |
| **Oral** | **Placebo** | 10 | 7 | 70.0 | 0.40 | 8 | 80.0 | 0.60 |
| **Oral** | **5 mg THC** | 10 | 5 | 50.0 | 0.00 | 6 | 60.0 | 0.20 |
| **Oral** | **10 mg THC** | 10 | 5 | 50.0 | 0.00 | 6 | 60.0 | 0.20 |
| **Vaporized** | **Placebo** | 9 | 5 | 55.6 | 0.11 | 5 | 55.6 | 0.11 |
| **Vaporized** | **2 mg THC** | 9 | 5 | 55.6 | 0.11 | 5 | 55.6 | 0.11 |
| **Vaporized** | **4 mg THC** | 10 | 8 | 80.0 | 0.60 | 8 | 80.0 | 0.60 |

**S15. Pharmacodynamic Effect-Size Summary**
**Table S6. Magnitude of all pharmacodynamic effects by route and dose (peak effect vs placebo), including non-significant outcomes.**

| **Pharmacodynamic Effect-Size Summary** | | | | | | | | |
| --- | --- | --- | --- | --- | --- | --- | --- | --- |
| **Domain** | **Outcome** | **Route** | **Dose** | **Peak time** | **MD [95% CI]** | **Hedges' g** | **p** | **FDR** |
| **Subjective** | Pleasurable (VAS) | Oral | 5 mg | 150 min | +19.1 [+3.4, +34.8] | +0.84 | .021 | .051 |
|  |  |  | 10 mg | 90 min | +22.8 [+2.0, +43.6] | +0.77 | .034 | .059 |
|  |  | Vaporized | 2 mg | 30 min | +21.0 [-9.1, +51.0] | +0.68 | .153 | .183 |
|  |  |  | 4 mg | 30 min | +35.1 [+12.2, +58.1] | +1.49 | .007 | .021 |
|  | Stimulatory (VAS) | Oral | 5 mg | 150 min | +25.3 [+9.0, +41.5] | +1.19 | .007 | .021 |
|  |  |  | 10 mg | 120 min | +33.3 [+15.3, +51.3] | +1.41 | .002 | .013 |
|  |  | Vaporized | 2 mg | 5 min | +19.0 [-5.4, +43.4] | +0.73 | .114 | .152 |
|  |  |  | 4 mg | 5 min | +42.0 [+20.7, +63.3] | +1.76 | .001 | .013 |
|  | Aversive (VAS) | Oral | 5 mg | 60 min | +6.6 [+0.1, +13.1] | +0.68 | .046 | .069 |
|  |  |  | 10 mg | 360 min | +1.7 [-3.1, +6.5] | +0.24 | .459 | .459 |
|  |  | Vaporized | 2 mg | 5 min | +3.3 [-2.7, +9.3] | +0.50 | .234 | .255 |
|  |  |  | 4 mg | 60 min | +10.5 [+1.3, +19.7] | +0.91 | .028 | .056 |
| **Reinforcement** | MCP crossover (USD) | Oral | 5 mg | — | -0.4 [-7.5, +6.7] | -0.04 | .911 | .911 |
|  |  |  | 10 mg | — | +2.1 [-4.9, +9.1] | +0.21 | .533 | .710 |
|  |  | Vaporized | 2 mg | — | +4.8 [-0.1, +9.7] | +0.70 | .052 | .104 |
|  |  |  | 4 mg | — | +9.0 [+3.6, +14.5] | +1.05 | .004 | .015 |
| **Cognition** | HVLT-R Total (words) | Oral | 5 mg | — | +0.9 [-3.2, +5.0] | +0.14 | .644 | .943 |
|  |  |  | 10 mg | — | -1.3 [-5.4, +2.7] | -0.22 | .489 | .943 |
|  |  | Vaporized | 2 mg | — | +2.2 [-4.4, +8.7] | +0.36 | .475 | .943 |
|  |  |  | 4 mg | — | +1.2 [-3.3, +5.8] | +0.28 | .570 | .943 |
|  | HVLT-R Delayed (words) | Oral | 5 mg | — | +1.0 [-0.4, +2.4] | +0.33 | .139 | .943 |
|  |  |  | 10 mg | — | +0.2 [-1.2, +1.6] | +0.06 | .755 | .943 |
|  |  | Vaporized | 2 mg | — | +0.8 [-1.7, +3.2] | +0.32 | .484 | .943 |
|  |  |  | 4 mg | — | +0.1 [-1.4, +1.7] | +0.07 | .867 | .943 |
|  | CPT throughput | Oral | 5 mg | — | -2.2 [-17.6, +13.1] | -0.11 | .757 | .943 |
|  |  |  | 10 mg | — | -0.5 [-15.8, +14.8] | -0.02 | .943 | .943 |
|  |  | Vaporized | 2 mg | — | +2.7 [-9.8, +15.3] | +0.12 | .642 | .943 |
|  |  |  | 4 mg | — | -1.1 [-12.7, +10.6] | -0.06 | .842 | .943 |
| **Cardiovascular** | Systolic BP (mmHg) | Oral | 5 mg | 360 min | -0.8 [-10.9, +9.2] | -0.07 | .857 | .956 |
|  |  |  | 10 mg | 360 min | +0.4 [-9.5, +10.2] | +0.03 | .940 | .956 |
|  |  | Vaporized | 2 mg | 5 min | -1.1 [-8.2, +6.1] | -0.12 | .750 | .956 |
|  |  |  | 4 mg | 3 min | -8.1 [-18.7, +2.5] | -0.61 | .118 | .497 |
|  | Diastolic BP (mmHg) | Oral | 5 mg | 120 min | +0.3 [-6.4, +7.0] | +0.03 | .927 | .956 |
|  |  |  | 10 mg | 240 min | +0.6 [-5.2, +6.4] | +0.08 | .831 | .956 |
|  |  | Vaporized | 2 mg | 3 min | -5.3 [-12.2, +1.7] | -0.77 | .124 | .497 |
|  |  |  | 4 mg | 5 min | -5.9 [-15.2, +3.4] | -0.59 | .192 | .575 |
|  | Heart rate (BPM) | Oral | 5 mg | 5 min | +0.2 [-7.1, +7.4] | +0.02 | .956 | .956 |
|  |  |  | 10 mg | 360 min | +0.5 [-6.7, +7.7] | +0.04 | .884 | .956 |
|  |  | Vaporized | 2 mg | 15 min | +1.0 [-6.6, +8.5] | +0.12 | .780 | .956 |
|  |  |  | 4 mg | 5 min | +11.6 [+3.7, +19.5] | +1.35 | .009 | .106 |

*All active-dose contrasts versus placebo. For DEQ and cardiovascular outcomes the value is the peak change from the pre-dose baseline (peak time shown); reinforcement and cognition are single per-session measures. MD = mean difference (active minus placebo), native units, 95% CI; Hedges' g is the mean difference divided by the average of the model-estimated dose-specific SDs; + = increase vs placebo. FDR = Benjamini-Hochberg false-discovery-rate-adjusted p-value, computed within each outcome domain (subjective, reinforcement, cognitive, cardiovascular) across all active-dose contrasts shown; adjusted p < .05 indicates the contrast survives correction for multiple comparisons.*

**Table S7. Omnibus dose tests by outcome and route.**

| **Main Dose Tests** | | | | | |
| --- | --- | --- | --- | --- | --- |
| **Domain** | **Outcome** | **Route** | **F (df)** | **p** | **FDR** |
| **Subjective** | Pleasurable (VAS) | Oral | 4.31 (2, 12.2) | .038 | .058 |
|  |  | Vaporized | 5.95 (2, 10.6) | .019 | .037 |
|  | Stimulatory (VAS) | Oral | 10.47 (2, 9.8) | .004 | .013 |
|  |  | Vaporized | 9.32 (2, 10.9) | .004 | .013 |
|  | Aversive (VAS) | Oral | 2.41 (2, 10.9) | .136 | .136 |
|  |  | Vaporized | 2.98 (2, 9.2) | .100 | .120 |
| **Reinforcement** | MCP crossover (USD) | Oral | 0.41 (2, 12.2) | .670 | .670 |
|  |  | Vaporized | 6.22 (2, 9.8) | .018 | .036 |
| **Cognition** | HVLT-R Total (words) | Oral | 1.28 (2, 12.4) | .312 | .936 |
|  |  | Vaporized | 0.30 (2, 10.4) | .745 | .942 |
|  | HVLT-R Delayed (words) | Oral | 1.73 (2, 13.1) | .215 | .936 |
|  |  | Vaporized | 0.27 (2, 9.6) | .772 | .942 |
|  | CPT throughput | Oral | 0.06 (2, 11.2) | .946 | .946 |
|  |  | Vaporized | 0.25 (2, 9.7) | .785 | .942 |
| **Cardiovascular** | Systolic BP (mmHg) | Oral | 0.04 (2, 11.0) | .958 | .987 |
|  |  | Vaporized | 1.45 (2, 10.3) | .278 | .556 |
|  | Diastolic BP (mmHg) | Oral | 0.02 (2, 11.6) | .977 | .987 |
|  |  | Vaporized | 1.54 (2, 10.4) | .259 | .556 |
|  | Heart rate (BPM) | Oral | 0.01 (2, 12.4) | .987 | .987 |
|  |  | Vaporized | 8.82 (2, 10.3) | .006 | .035 |
| *Omnibus dose tests from the linear mixed models (2 numerator degrees of freedom; Kenward-Roger denominator degrees of freedom). FDR = Benjamini-Hochberg false-discovery-rate-adjusted p-value, computed within each outcome domain (subjective, reinforcement, cognitive, cardiovascular) across the omnibus tests in that domain; adjusted p < .05 indicates the effect survives correction for multiple comparisons. Omnibus dose effects for stimulatory ratings in both arms, vaporized pleasurable ratings, vaporized reinforcement, and vaporized heart rate survived correction.* | | | | | |

**S16. Within-Participant Exposure–Response**

We related each pharmacodynamic outcome's peak post-dose response to peak total active exposure within participants (Methods, Section 2.10). Total active concentration was defined as C_total = [THC] + S x [11-OH-THC], where S is the weight on the active metabolite 11-OH-THC relative to parent THC; the primary analysis used S = 1 (equal potency). Models were linear mixed-effects regressions of the raw peak response on exposure, with the pre-dose baseline as a covariate and a random intercept per participant; effect sizes are within-participant partial correlations (r). Reinforcement and cognition were assessed once per session, without a pre-dose baseline, and were therefore not amenable to this baseline-adjusted within-participant model. These associations were robust to the metabolite weighting: within-participant r was essentially unchanged as S varied from 0 (metabolite ignored) to 2 (twice the parent), and the vaporized arm was invariant given its negligible 11-OH-THC.

**Table S8. Within-participant exposure-response by outcome and route.** Within-participant partial correlation (r) between the peak post-dose response and the peak total active concentration (Cmax of THC plus 11-OH-THC), from a linear mixed-effects model with the pre-dose baseline as a covariate and a random intercept per participant. r (Cmax) is the primary metric; r (AUC) repeats the model with total exposure (AUC 0 to 8 h) as a sensitivity. Cells shaded by r (red = positive, green = negative; deeper = stronger), significant primary associations in bold.

| **Within-Participant Exposure–Response** | | | | | | |
| --- | --- | --- | --- | --- | --- | --- |
| **Domain** | **Outcome** | **Arm** | **r (Cmax)** | **p (Cmax)** | **r (AUC)** | **p (AUC)** |
| **Subjective (DEQ)** | DEQ Pleasurable | Oral | **+0.47** | .019 | +0.32 | .116 |
|  | DEQ Pleasurable | Vaporized | **+0.65** | <.001 | +0.39 | .045 |
|  | DEQ Stimulatory | Oral | **+0.64** | .001 | +0.51 | .007 |
|  | DEQ Stimulatory | Vaporized | **+0.71** | <.001 | +0.32 | .105 |
|  | DEQ Aversive | Oral | +0.00 | .993 | +0.05 | .809 |
|  | DEQ Aversive | Vaporized | +0.26 | .230 | +0.21 | .359 |
| **Cardiovascular** | Systolic BP | Oral | -0.08 | .656 | -0.02 | .898 |
|  | Systolic BP | Vaporized | +0.08 | .668 | -0.08 | .679 |
|  | Diastolic BP | Oral | +0.06 | .764 | +0.11 | .533 |
|  | Diastolic BP | Vaporized | -0.01 | .951 | +0.09 | .711 |
|  | Heart rate | Oral | -0.23 | .273 | -0.09 | .660 |
|  | Heart rate | Vaporized | **+0.65** | .001 | +0.56 | .003 |

**S17. Plasma THC Exposure in Relation to Body Weight (Per-Dose)**

We examined whether plasma THC exposure differed between participants according to body weight, for parent THC and its active metabolite 11-hydroxy-THC. Plasma exposure (peak concentration [Cmax] and area under the curve from 0 to 8 hours [AUC0–8h]) was related to body weight using Spearman rank correlations, separately at each active dose within each route (**Table S9**).

Oral exposure decreased with greater body weight, an association that reached significance at the higher oral dose (10 mg) for THC Cmax (ρ = -0.73, p = .021), 11-OH-THC Cmax (ρ = -0.75, p = .018), and 11-OH-THC AUC0–8h (ρ = -0.65, p = .049); at 5 mg the same correlations were weaker and non-significant (ρ = +0.00 to -0.39). Vaporized exposure showed no significant association with body weight at either dose. The pattern is consistent with a fixed oral dose producing lower plasma concentrations in heavier participants.

**Table S9. Plasma Exposure vs Body Weight.** Per-dose between-participant correlations of plasma exposure with body weight, by analyte, metric, and route. Values are the Spearman rank correlation (ρ) and its p between each participant's exposure (Cmax or AUC0–8h) and body weight at each active dose. Cells shaded by ρ (green = positive, red = negative; deeper = stronger); nominally significant associations (p < .05) in bold.

| **Plasma Exposure vs Body Weight** | | | **5 mg Oral / 2 mg Vaporized** | | **10 mg Oral / 4 mg Vaporized** | |
| --- | --- | --- | --- | --- | --- | --- |
| **Analyte** | **Metric** | **Route** | **ρ** | **p** | **ρ** | **p** |
| **THC** | Cmax | Oral | +0.00 | 1.000 | **-0.73** | **.021** |
|  |  | Vaporized | -0.28 | .472 | -0.28 | .434 |
|  | AUC0–8h | Oral | -0.22 | .537 | -0.53 | .123 |
|  |  | Vaporized | -0.36 | .389 | -0.11 | .763 |
| **11-OH-THC** | Cmax | Oral | -0.30 | .407 | **-0.75** | **.018** |
|  |  | Vaporized | -0.12 | .761 | -0.43 | .211 |
|  | AUC0–8h | Oral | -0.39 | .263 | **-0.65** | **.049** |
|  |  | Vaporized | -0.22 | .604 | +0.12 | .738 |
